# Predicting Worry Mental States using Long Short-Term Memory (LSTM) Recurrent Deep Neural Networks

**DOI:** 10.64898/2026.08.14.26360462

**Authors:** J.Y. Campion, T. Desmidt, J.J. Gross, D.L. Tudorascu, C. Andreescu, H.T. Karim

**Author notes:** **Corresponding Author(s):** Helmet T. Karim, PhD, Assistant Professor of Psychiatry and Bioengineering, University of Pittsburgh, Carmen Andreescu, MD, Professor of Psychiatry and Bioengineering, University of Pittsburgh.

## Abstract

Severe worry is a transdiagnostic syndrome associated with significant morbidity in older adults. In this study, we aim to infer worry-related mental states though brain activity timeseries. We acquired fMRI on two cohorts (N=116 and N=88), using an in-scanner worry induction and reappraisal task. We trained a recurrent long short-term memory (LSTM) neural network, using the first cohort as the train/validation and the second cohort as an independent test set. We predicted worry induction, reappraisal, and neutral states (area under the curve 0.89, 0.77, 0.91 for the test set and 0.78, 0.63, 0.81 for the independent set). The model was most accurate when participants reported high worry during the induction state. Dorsal attention network, and networks seeded on the anterior hippocampus, and supplementary motor area were most important for predicting worry states. The LSTM approach may have critical translational implications for identifying and treating severe worry in older adults.

---

Worry is a complex affective and cognitive process, which – in mild forms – may confer evolutionary adaptative advantages^1^. In the severe forms encountered in mood and anxiety disorders, worry is difficult to control, associated with negative affect, decreased quality of life, and significant morbidity^1,2^. In older adults, severe worry has been associated with cardiovascular and cerebrovascular disease^3^ ^4^, cognitive impairment, heightened risk of dementia, and accelerated brain aging^5–9^. Severe worry is highly prevalent in older adults (20% prevalence in community samples^10^), but frequently undertreated. This may be related to difficulties in identifying severe worry in older adults, due to generational cultural differences that lead older adults to perceive severe worry as a weakness and focus instead on reporting associated somatic symptoms^11^. Given the weight of self-reported symptoms in mental health diagnoses, these age-related challenges may interfere with illness recognition and treatment, and may worsen long-term vascular and cognitive health. Thus, individual prediction of mental states may advance our understanding of severe worry in older adults and may lead to new and more effective interventions.

There is a paucity of studies exploring neural markers of severe worry. The few available studies do not allow for definitive conclusions due to variations in design and sample characteristics^12^, including categorical approaches focusing on Generalized Anxiety Disorder (GAD)^12–14^ versus dimensional approaches focusing on worry severity. Our group has previously designed a naturalistic in-scanner worry induction and reappraisal task that was validated on several independent, age-varied cohorts^13,15,16^. One of our early studies using this task described different substrates for initiation, maintenance, and severity of worry^17^ involving anterior cingulate cortex (ACC), supplementary motor area (SMA), visual cortex, caudate, thalamus, insula, parahippocampus, hippocampus and amygdala, showing altered cerebral blood flow associated with severe worry. More recently, these results were subsequently replicated and expanded using BOLD fMRI acquisitions in a larger cohort^16^. The previous studies published by our group^13,16,17^ used traditional group level statistics that entail the decreased sensitivity, multiple comparisons, and partial modelization of the spatiotemporal complexity of worry processes. Deep learning models may potentially expand on our understanding of severe worry while accounting for individual prediction and variability.

In this study, we aimed to train a deep learning model using task-based blood oxygenation level-depent (BOLD) activation patterns to predict the specific states of worry induction and reappraisal [worry, neutral, reappraisal states]. This model learned from timeseries data from various regions to predict the above states per participant. Advances in deep learning approaches – like long short-term memory (LSTM) networks – allow a more nuanced analysis of worry states by analyzing the personalized temporal evolution of neural activity patterns. This technological advancement allows for more individualized understanding of how worry-related brain states fluctuate over time and interact with other cognitive and emotional processes.

To achieve this aim, we analyzed timeseries data from the in-scanner worry induction and reappraisal task from two independent cohorts of older adults. We hypothesized that LSTM networks could effectively identify and predict the temporal dynamics of worry and reappraisal-related neural states using regional brain activity timeseries. We evaluated the relative importance of regions and networks on predicting worry states. We also evaluated the temporal dynamics of the brain networks associated with worry induction.

## Results

### Population Description

We used two independent cohorts from R01 MH108509-05 (Functional Neuroanatomy Correlates of Worry in Older Adults [FINA], N= 116) and R01 MH 108509-06 (The RAW Brain – The Effect of Rumination, Anxiety and Worry on Aging and Dementia Risk [RAW], N = 88). Clinical and demographic characteristics are presented in Supplement Table 1. The train and validation FINA samples had significantly higher levels of anxiety, worry and rumination compared to the RAW sample. All three samples (train FINA, validation FINA, RAW) had a larger proportion of women than men.

### Model Performance

We trained an LSTM to predict worry, neutral, and reappraisal states on regional timeseries data from the training set from FINA, validated using the other part of FINA and tested its accuracy on RAW. Our model predicted values for worry, neutral, and reappraisal states from probability of 0 to 1 for each second. A value of 1 was associated with high likelihood to be in one of the three states. We computed a Mean Absolute Error (MAE) and the area under the curve (AUC) of the receiver operating characteristic (ROC) curve as accuracy metrics for each condition. The MAE was 0.0712 in the FINA validation and 0.1275 in the RAW independent dataset. We report in Supplement Figure 1 the MAE when the model was predicting using a random 20 second length chunk versus task-synchronized chunks (as compared to the whole sequence). The overall AUC of worry, neutral, and reappraisal states was 0.89, 0.77, 0.91 in the FINA validation and 0.78, 0.63, 0.81 in the RAW dataset, respectively. To highlight variability in classification performance across participants, we estimated AUC per participant (Figure 1). In summary, our model performed accurately and we show that this model generalizes to an independent test set; and the accuracy of the model was best on the worry and neutral states but had difficulty differentiating worry from reappraisal.

**Figure 1.**
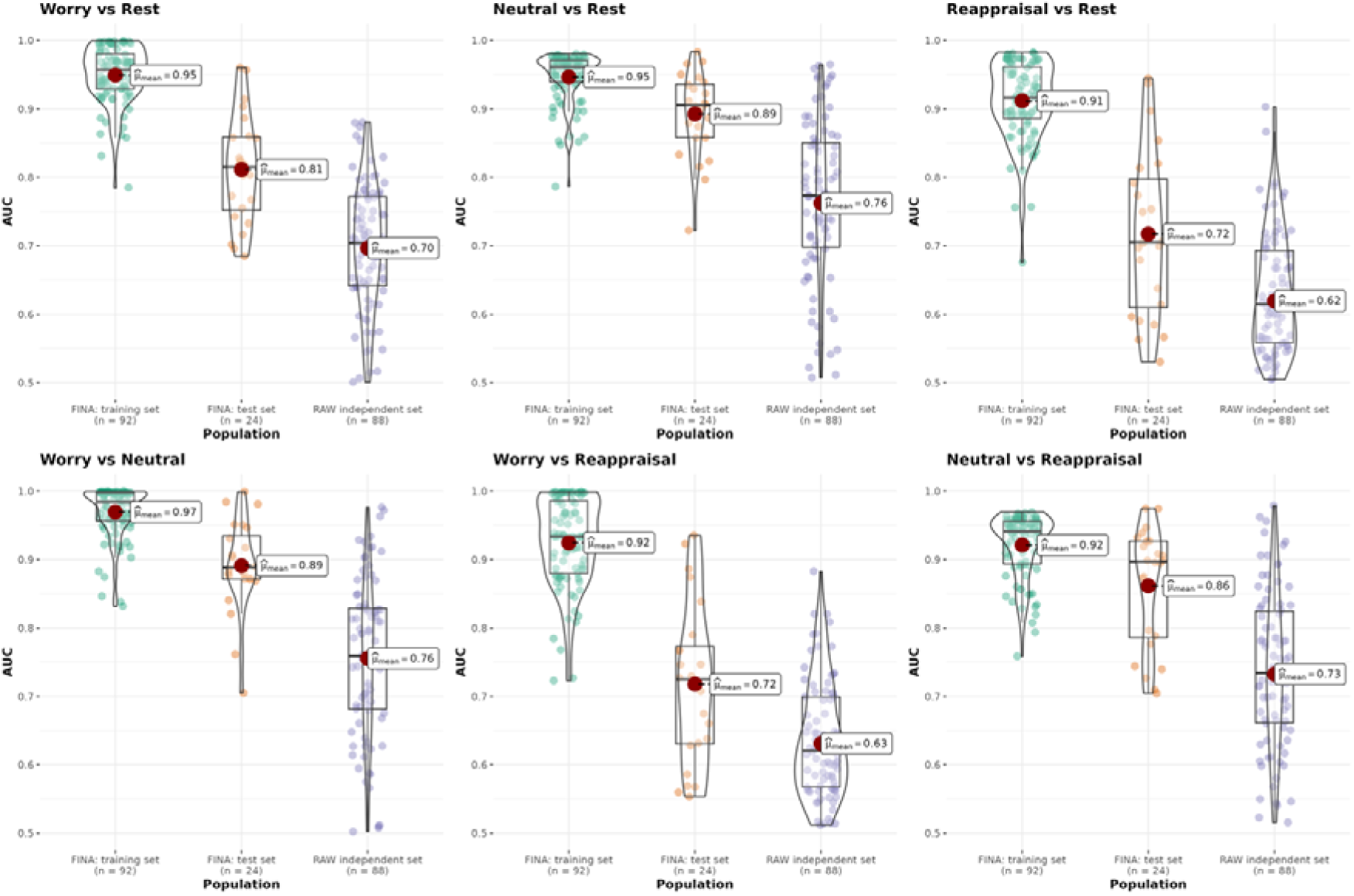
Model performance for each subset expressed as AUC ROC for different classification task based on 25s task synchronized sequences. In this plot, we perform AUC ROC for classification of each task against the rest and paired classification of tasks. Each of these metrics are performed per participant. The mean performance is indicated by the red dot. A boxplot and violin plot is added to show the distribution. Significant differences between subsets after Holm-Bonferroni correction are reported. Overall, these figures show a loss of performances and more variability with the **FINA: test set** and **RAW independent set**.

### Factors Associated With Performance

We wanted to evaluate whether this model was more or less accurate depending on various clinical and demographic factors (e.g. age, sex, worry severity, rumination severity). We tested this using a linear mixed effect model, evaluating associations between model accuracy and potential clinical, demographic factors, and condition (worry, neutral, and reappraisal). The RAW study included 20 participants from FINA who were re-tested 2-3 years later (13 from the training dataset and 7 from the validation dataset). Due to this, we added a factor to account for this set who were a part of the FINA. Each block was designed as follows: 10 seconds of personalized statement worry induction and 15 seconds of maintenance, followed by worry severity rating from 1 (no worry) to 5 (severe worry). The scaled rating is computed from this metric. All results are shown in Figure 2.

**Figure 2.**
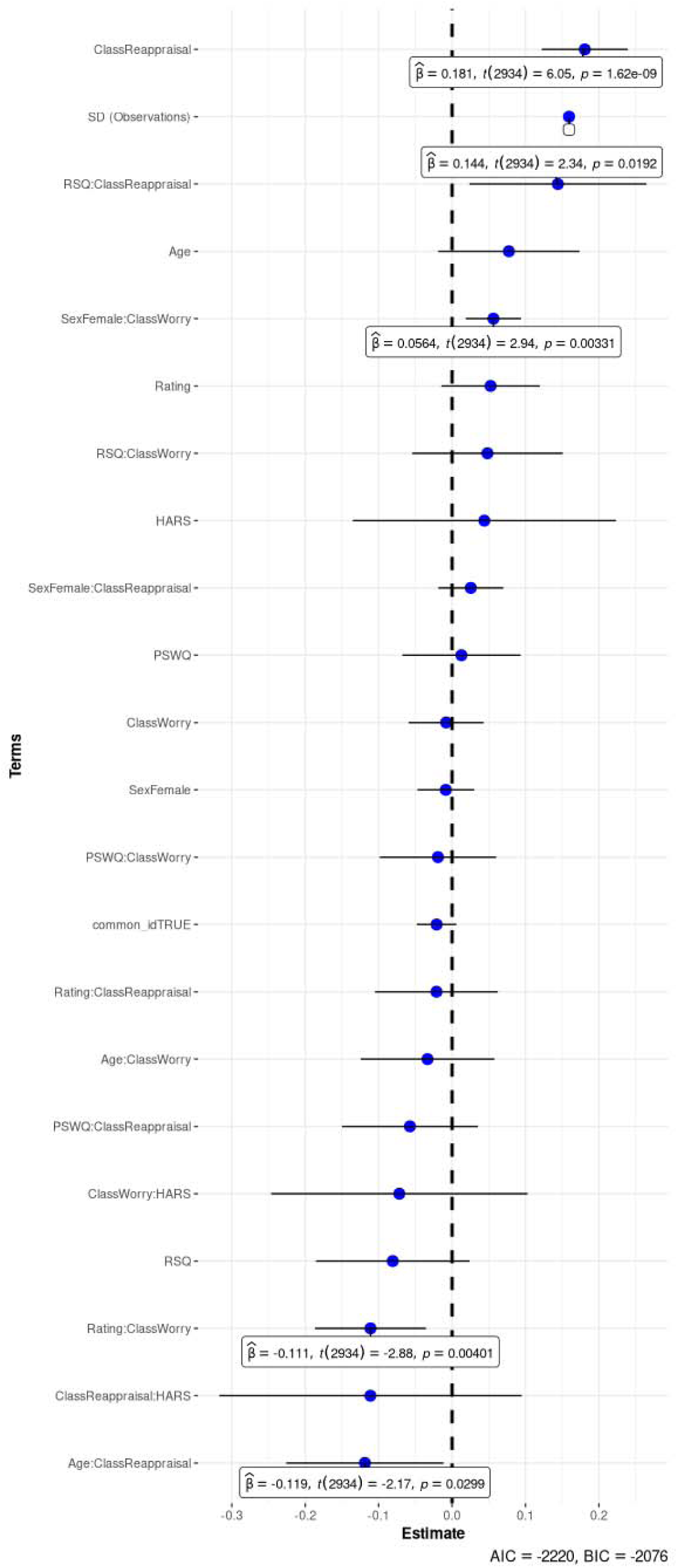
Linear Mixed Effect model coefficients affecting absolute error (performance). The model is a linear mixed effect model with subject as a random effect, predicting the absolute error and using age, sex, worry rating (the severity of the worry induced), condition (worry, neutral or reappraisal), common_id (represent if the subject was recruited in FINA and RAW at two different sessions), RSQ, PSWQ, HARS and interactions between class and other variables. All significant coefficients are displayed with caption reporting beta and p values. AIC and BIC are reported on the bottom right corner. For example, higher ß values for reappraisal show that reappraisal (compared to neutral) was associated with a higher absolute error or less accurate prediction.

The model’s performance was stronger for predicting worry states and lower for predicting reappraisal states. When predicting reappraisal, the model was less accurate in individuals with higher rumination (as measured by RSQ-Rumination Subscale) and those who were younger compared to those who were older. When predicting worry, the model was less accurate when participants rated the block as inducing lower worry severity compared to when they rated that the worry block induced high worry severity. In addition, when predicting worry, the model was less accurate in women compared to men.

### Model’s Importance Over Time

We computed the mean of absolute values of SHapley Additive exPlanations value (smaShap, min-max scaled) for each timepoint – this identified the most important timepoints for identifying worry, neutral, and reappraisal blocks. We plot importance over time in Figure 3 with peaks around the 8-10^th^, 15-16^th^ and 25^th^ seconds. We found that the neutral condition showed less contrast compared to the other conditions.

**Figure 3.**
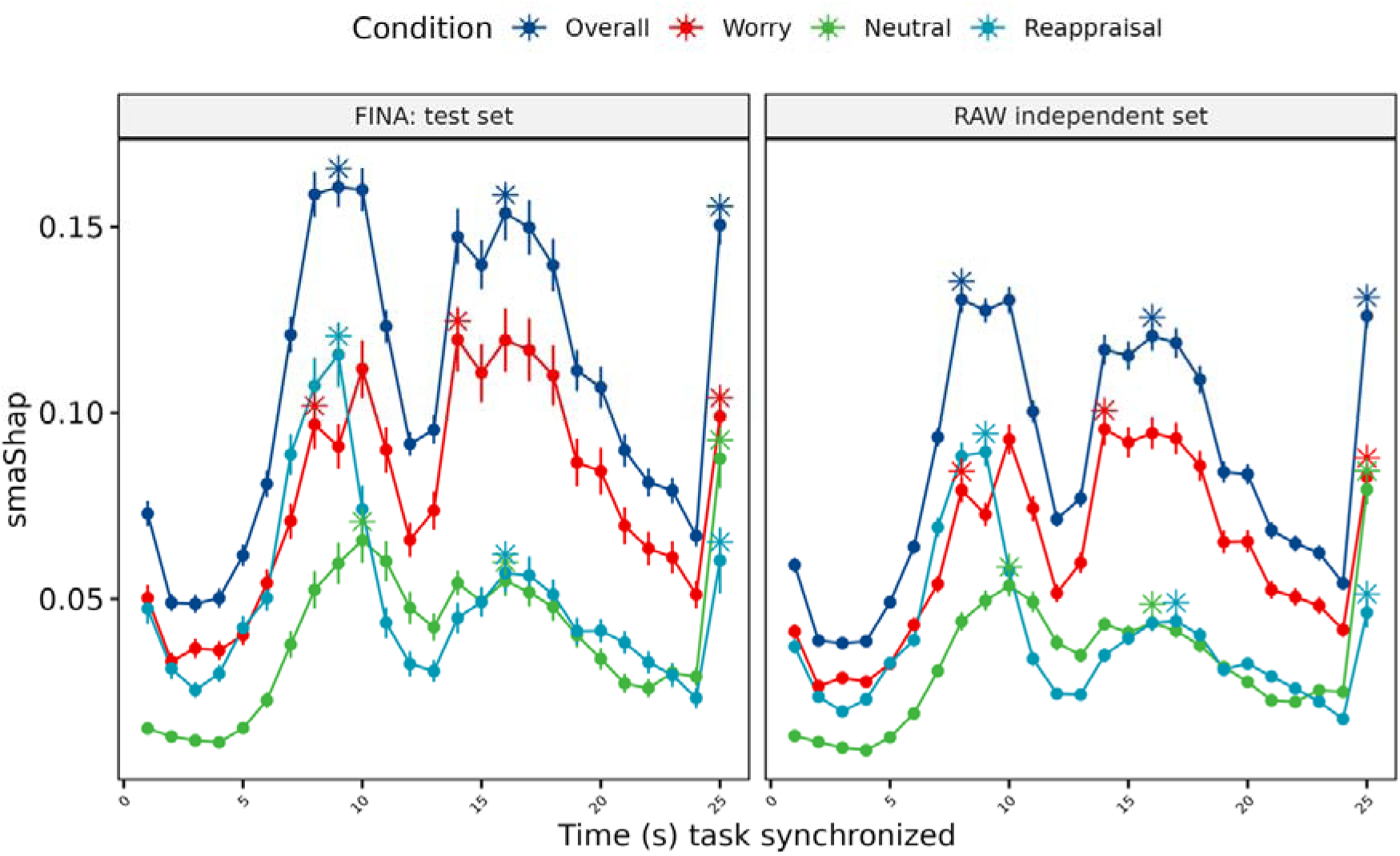
Mean absolute Shap value (scaled) per condition across a block, stars are local maxima. Mean absolute values of Shap values (smaShap, scaled) are plotted across the block for each condition worry, neutral and reappraisal. We plot the overall mean as well. Stars indicate local maxima of Shap values. We show that there are local maxima around 9s ± 1s, 15s ± 1s, and the 25s timepoint of the block for each task.

### Model’s Importance Across Networks

We also computed SHAP values to understand which regions of interest (ROIs) were most important. To improve interpretability of these values, we computed a network importance measure by computing the dot product between L2 regularized ROI importance for the model and different reference networks as defined by Smith et al. networks^18^. In addition, we evaluated their relative importance over time. These plots are shown in Figure 4 with the Smith networks visualized. The most important network was defined as those that have at minimum 10% of timepoints with the highest Shap value.

**Figure 4.**
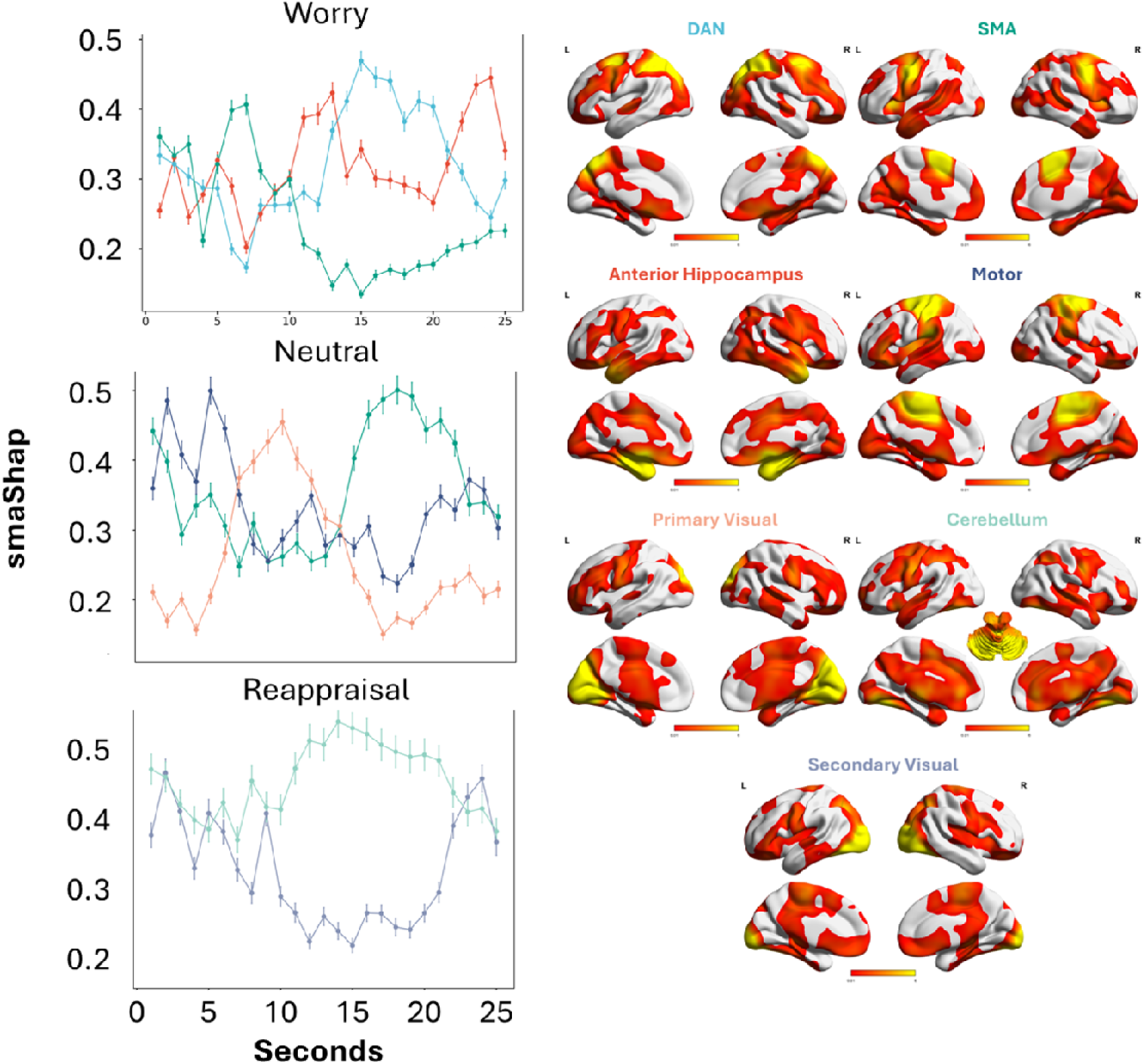
Network importance over time plotting mean absolute Shap (scaled) Mean absolute of Shap values (smaShap, scaled) are plotted for each network for each condition. This was defined as networks that have at minimum 10% of timepoints with the highest Shap value. We plot the networks smaShap values over time with means and 95% confidence intervals. We also plot the relevant networks in Brain Net viewer.

The most important network for predicting worry was the dorsal attention network (DAN). Temporally, we found that the SMA network was most important for initiating worry; while the anterior hippocampus network and DAN were most important in the maintenance; and the anterior hippocampus was most important for the termination of worry. We plot all of the networks over time for worry in Supplement Figure 2.

The most important network for predicting neutral states was the SMA network. For neutral, the SMA and motor cortex networks were most important in the beginning; primary visual cortex was most important in the middle of the block; and SMA was most important at the end of the neutral block. We plot all of the networks over time for neutral in Supplement Figure 3.

The most important network for predicting reappraisal was the cerebellum network. The cerebellum was important across time, while the secondary visual network was most important at the beginning and end of the reappraisal blocks. We plot all of the networks over time for reappraisal in Supplement Figure 4.

### Exploratory: Factors Influencing the SMA Importance for the Model

We found that the SMA network was one of the most important networks for both worry and neutral conditions. We wanted to evaluate what clinical and demographic factors were associated with SMA importance using a linear mixed effects model. The subject identifier was used as random effect, an intercept was set for timepoints and we used the same factors as the previous model except the FINA-RAW cross inclusion. The results are reported in Figure 5.

**Figure 5.**
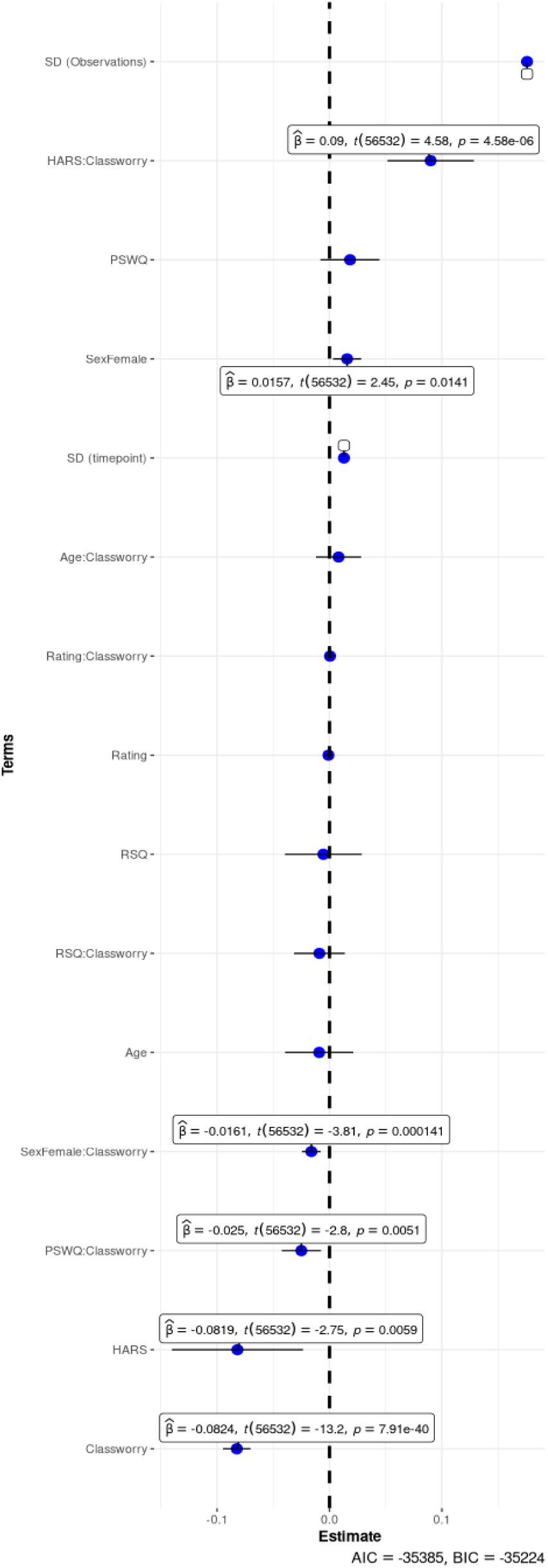
Linear mixed effect model predicting SMA Shap values. The model is a linear mixed effect model with subjects as random effect, predicting the mean absolute Shap values of Supplementary Motor Area (smaShap SMA, scaled) and using age, sex, rating, class (worry or neutral), RSQ, PSWQ, HARS and interactions between class and other variables. All significant coefficients are displayed with caption reporting ß and p values. AIC and BIC are reported on the lower right corner.

While predicting worry (relative to neutral), we found that the SMA was more important in individuals with greater anxiety (HARS), lower worry (PSWQ), and who were male (compared to female). All the coefficients were reported in Figure 5.

## Discussion

We designed an artificial neural network trained to decode worry, neutral, and reappraisal states based solely on task-based fMRI timeseries data without clinical input. The model’s performance remained stable across datasets, scanner type (3T vs 7T), and largely across clinical presentations. Using Shap values, we showed the spatiotemporal contribution of various regions and networks for both worry induction and reappraisal of worry. This pattern of activity replicates our past work focused on the networks involved in the induction, maintenance, and severity of worry processes^16, 17^. This type of decoding may also help identify worry states during resting state acquisitions which are more ubiquitous and can be applied with lower participant burden.

Previous studies have used fMRI timeseries data combined with deep neural networks to decode tasks. Wang et al. used Human Connectome Data to differentiate seven different tasks using a deep neural network with high accuracy (89-95% accuracy) but they predicted the task (working memory, gambling, language task, etc.)^19^ similar to approaches implemented by Kucukosmanoglu et al.^20^ who used longitudinal data to predict tasks. These studies did not predict task states during the task, but rather the task label. Similar to our approach, Thomas et al. used 100 participants’ data to predict different stimulus from an N-back task showing four different stimuli (body, face, place and tool), by using a voxel-wise and layer-wise LSTM to predict these with moderate accuracy (∼68%) on a small held out test set (n=30)^21^. We were able to accurately identify worry and neutral states with high accuracy (AUC 0.78-0.81) and reappraisal with moderate accuracy (AUC 0.63) using timeseries task fMRI data and testing on a large independent test set.

We found that the model was less accurate at decoding reappraisal compared to neutral and worry, and that this was even less accurate in individuals with high rumination and who were younger in this sample (e.g., in their 50s). The lower accuracy for reappraisal may be due to interindividual variability in cognitive reappraisal effectiveness and processes^22,23^. Individuals with high rumination may have more difficulty reducing negative emotional arousal through reappraisal^24–26^ and older adults experience less rumination compared to younger adults^27,28^ – this may explain why our model was more accurate in older individuals with lower rumination. We found that the model was less accurate at decoding worry compared to neutral and reappraisal when individuals rated that block as inducing lower worry. This may be explained by blurred boundaries between low worry and neutral, e.g., low worry states may be more likely to be labeled as ‘neutral’. In addition, we found that the model was less accurate in women compared to men when decoding worry. In part, this may be due to recruitment – we had more female participants and potentially more variability to capture. It may also be due to differences in neural activity during worry – this may be reflected in both differences in brain activity induced during worry between men and women but also in their prevalence. In one study exploring anxiety inducing stressful imagery, women showed greater activation in regions involved in emotion regulation and arousal inhibition while men show greater activation in the caudate, cingulate, thalamus, and cerebellum^29^. Finally, women experience anxiety and worry at a higher rate compared to men – this may partly be related to this effect^30^.

Our analysis evaluating model importance used Shap values to provide critical insights into the spatio-temporal importance for prediction. The analysis of time attention revealed that specific timepoints contributed disproportionately to the model’s prediction performance. These generally aligned with the midpoint of the blocks (8-11sec and 15-16sec) as well as the end of the block. These may reflect the maintenance processes of worry^17^ and potentially “meta” processes associated with rating.

We also evaluated model importance across networks and time – we found the relative importance of certain regions and networks in predicting worry, neutral, and reappraisal states. We found that SMA network, DAN and anterior hippocampus networks were critical for predicting the initiation, maintenance, and termination of worry blocks, respectively. Of note, the SMA network included parts of the default mode and attention networks while anterior hippocampus network primarily included parts of the limbic cortex including amygdala, subgenual anterior cingulate, and parts of the basal ganglia. These results confirm our previous worry induction model, describing specific regions implicated in the initiation (supplemental motor, dorsal anterior cingulate, caudate/thalamus) and the maintenance (insular cortex, hippocampus, caudate/thalamus, and amgydala) of the worry process^16,17^. These networks have been implicated in recruitment of biased autobiographic memory that allows retrieving negative memories^31^ and helping context or scenario elaboration^31^. The DAN engagement supports features like disengagement difficulties and could support continued attention bias associated with difficulty to disengage^32–35^.

The reappraising state was predicted first and foremost by a cerebellum-centered network^14,36^ that included prefrontal cortex, part of basal ganglia and limbic regions, followed by the secondary visual cortex. The cerebellum and its associated prefrontal regions has been more recently implicated, through its limbic and prefrontal connections^22,37^, in top-down modulation of emotional responses and regulation processes. The secondary visual cortex could be interpreted as support for mental imagery that are engaged to support the reappraisal process^38,39^.

Motor network, the primary visual cortex, and the SMA network were associated with neutral state prediction. 78% of these neutral blocks were proceeded by a worry block and 22% by a reappraisal block. We can hypothetise that the tail of passive dissolution of worry or attempts at implicit emotion regulation engaging the SMA ^17,35,39–43^. The involvement of the primary visual cortex in the transition part of the block could be associated with the reading of neutral statements during this portion of the task.

Our study has several limitations, mostly associated with the datasets used for training the model. The worry distribution differed between the two studies, leading to an unbalanced representation of high and low worriers in train and test sets. Overall, the sample size was modest but allowed for independent testing. Future testing on larger samples is warranted. Also, we did not test this model in younger individuals so we can not generalize the model to younger populations. We trained this model on 3T and tested on 7T fMRI data which could have affected performance.

We trained a deep learning model that accurately predicted worry and neutral states per second using fMRI timeseries data. We tested this on an independent test set and evaluated feature importance to better characterize these complex brain dynamics. Once we validate this on larger datasets and on different mental states, and potentially trained on more widely available fMRI (e.g., resting state), this model may have important implications for understanding worry during more natural states – specifically by applying its prediction on resting state data. This could uncover more natural processes of worry and help improve our understanding of worry across larger populations without task-based worry induction data.

## Online Methods

### Participants and Study Design

We recruited participants from two different studies: “The Functional Neuroanatomy Correlates of Worry in Older Adults” (FINA, R01 MH 108509) and its competing continuating “The RAW brain - the effect of rumination, anxiety, and worry on aging and dementia risk” (RAW, R01 MH 108509-06). Participants in both studies (N=117 in FINA, N=88 in RAW) were recruited across a varying range of worry severity. Participants over 50 years or older with or without a categorical diagnosis of anxiety and/or depressive disorder were included in the studies. Participants with neurodevelopment disorders, intellectual development disorders, bipolar disorder, psychosis, personality disorder, and major neurocognitive disorder were excluded as well as those with increased suicide risk, history of drug/alcohol abuse within last 6 months, use of high dose of benzodiazepines (≥2mg lorazepam), uncorrected vision, below 6^th^ grade level of reading, and any contraindications of MRI. Low dose psychotropics were permitted for medical reasons like sleep disorders or pain. We recruited from the Pittsburgh area using Pitt+Me (an online resource at Pitt), in-person recommendations, flyers, radio/television advertisements and telephone scripts. Both studies were approved by the University of Pittsburgh Institutional Review Board and all participants gave written informed consent before participation. In the case of individuals who screened for mild cognitive impairment, capacity to consent was assessed prior to consent. The population decription is described in Supplement Table 1.

### Assessments

We collected demographic information (age, sex, race, education), and assessed the following: worry severity (PSWQ), overall anxiety severity (HARS, Hamilton Anxiety Rating Scale)^2^, rumination severity (RSQ, Rumination Subscale Questionnaire)^3^ and depression severity (MADRS, Montgomery-Asberg Depression Rating Scale)^4^. We also assessed and computed the brooding and reflection components of rumination from RSQ^6^. Cognitive status was evaluated using a comprehensive battery including Repeatable Battery for the Assessment of Neuropsychological Status (R-BANS)^40^, the sorting and verbal fluency tests of the Delis-Kaplan Executive Function Scale (DKEFS)^41^, IQCODE (interview with informant to determine change in function) and PASS (measure of instrumental skills in daily living)^42^. The presence or absence of MCI was established through adjudication by an expert neuropsychologist (MB).

### Worry Induction and Reappraisal Task

We have published extensively on this naturalistic task designed to elicit in-scanner personalized worry and reappraisal^30,33^. The full details of this task have been reported elsewhere^16^. In brief, prior to MR scans, we collected specific worry themes from each participant, who then generated both statements that would induce worry and statements that would reappraise worry. These conditions along with neutral statements, were shown in a block design with fixation blocks in between (Supplement Figure 5). Of note, the reappraisal blocks randomly followed half of the worry statements, in order to avoid anticipation. The task was shown in Psychtoolbox version 3 in MatLab. It is critical to note that while the interviewers helped the participants form these worry and reappraisal statements, they did not choose them, as these statements have to be relevant and effective for each participant. Each condition was shown for 25 seconds with inter-trial intervals of 10 seconds of a white fixation cross on a black background. Each participant received a version of the task that was pseudo randomly ordered to one of four sequences.

### Imaging

For the FINA study, MR scans were conducted at the MR Research Center at the University of Pittsburgh using a 3T Siemens MAGNETOM Prisma scanner and a 32-channel head coil. A sagittal, whole-brain T1-weighted magnetization prepared rapid gradient echo (MPRAGE) was collected with repetition time (TR)=2400ms, echo time (TE)=2.22ms, flip angle (FA)=8deg, field of view (FOV)=320×300 with 208 slices, 0.8mm^3^ isotropic resolution, 0.4mm slice gap, and GeneRalized Autocalibrating Partial Parallel Acquisition (GRAPPA) with acceleration factor of 2 (total time 6.63min). We collected 20 minutes of whole brain T2*-weighted blood oxygen level-dependent (BOLD) images using a gradient-echo echoplanar imaging sequence in axial orientation with TR=1000ms, TE=30ms, matrix size=96×96, voxel size=2.3mm^3^ isotropic (2.3mm slice gap), FA=45 degrees, and multiband acceleration of 5.

For the RAW study, we used a 7T Siemens MAGNETOM scanner with a custom tic-tac-toe coil. A sagittal, whole-brain T1-weighted magnetization prepared rapid gradient echo (MPRAGE) was collected with repetition time (TR)=3000ms, echo time (TE)=1.96ms, flip angle (FA)=8deg, field of view (FOV)=320×290 with 240 slices, 0.8mm^3^ isotropic resolution, 0.4mm slice gap, and GRAPPA with acceleration factor of 2. We collected 20 minutes of whole brain T2*-weighted blood oxygen level-dependent (BOLD) images using a gradient-echo echoplanar imaging sequence in axial orientation with TR=700ms, TE=20ms, matrix size=96×96, voxel size=2.0mm^3^ isotropic (2.0mm slice gap), FA=65 degrees, and multiband acceleration of 5.

### Preprocessing

Processing was conducted in statistical parametric mapping toolbox (SPM12)^10^. All interpolation was done with 4^th^ degree B-splines and similarity metrics were normalized mutual information. The MPRAGE was input into a segmentation to segment into 6 tissues^11^. We adjusted the number of Gaussians to two for white matter to account for white matter hypointensities. This segmentation generates a deformation field that can normalize functional images to a standard anatomic space (Montreal Neurological Institute or MNI space). We generated an intracranial volume mask by creating a single mask by thresholding the gray matter, white matter, and cerebrospinal fluid probability maps by 0.1 and then conducting image filling and image closing in MatLab. We applied this to the MPRAGE to generate a skull stripped MPRAGE.

For the functional images, we conducted motion correction. We then conducted skull stripping of the mean image and all functional images (using the brain extraction tool in FSL). We coregistered the mean skull-stripped functional image to the skull-stripped MPRAGE and applied this to all functional images. We normalized these files into MNI space (2mm^3^ isotropic resolution) using the structural deformation field and conducted spatial smoothing (full-width at half-maximum of 8mm).

We extracted Region of Interest (ROI) average time series with the cortical regions of Schaeffer atlas with 400 ROI and 81 subcortical and cerebellar regions of Automated Atlas Labelling version 3 (AAL3)^43,44^. Using the design of the study, we convolved the boxcars for the worry, neutral, and reappraisal blocks with the hemodynamic response function (HRF). These HRF convolved timeseries represent the expected brain activity for each task and were the variables that we predicted using regional activity.

### Network Analysis

In order to better understand the high dimensionality of the results, we used network level maps created using fMRI activity that were generated using independent components analysis – this generated 18 networks based on 30,000 healthy participants fMRI data by Smith et al ^45^. Note that these maps are simply named but represent complex regions – for example, the cerebellum network includes regions of the prefrontal cortex, basal ganglia, and default mode. We extracted the mean contribution for each ROI previously used. We considered the weights of each network as a vector and L2 regularzed these. The Shap and fMRI extracted values were averaged following the objectives and considered as well and regularized. After regularizing these vectors, we computed the dot product to estimate the contribution of each region and its association with each network – higher values indicate greater importance for that network.

### Train, Validation, and Test

The dataset was divided into 3 subsets: train, validation, and test. The test group only included data from RAW acquired with 7T MRI. The train and validation used data from FINA with 3T MRI and split with an 80% to 20% train-validation split. If needed, sequences were resampled using a cubic spline interpolation method to reflect a one second time resolution. If any sequence contained missing values, we excluded the participant thus we had a total of 116 participants in FINA and RAW, respectively. We excluded one FINA participant and twenty-five participants in RAW. Once subsets were defined, we randomly sampled the 1,200s length sequence into one hundred overlapping 20s samples per participant (excluding the 10 first seconds of acquisition).

### Model Architecture

In order to create our model, we incrementely increased its complexity using a test-driven development or build-measure-learn approach. We started with a bidirectional LSTM block followed by the decoding block, then added an another BiLSTM block, added an encoding block and finally added the recurrent connection. All these developments were first motivated by a maximum of learning and for the recurrent connection because of the underuse of the actual timepoint creating a smoothed prediction. Each step was tested and compared using different hyperparameters.

Our model was based on two recurrent bidirectional Long Short-Term Memory^46^ (BiLSTM) blocks, preceded by an encoding block and an output block. The model used hidden size as a parameter. The detailed parameters of each layer are described in Supplement Figure 6.

The encoding block started with a linear layer with a channel input of 481 and output of double hidden size. We then applied a ReLU, dropout and layer normalization. A second linear layer is applied with an output of hidden size and then ReLU applied.

The recurrent BiLSTM block was composed of two ways for input data. The first one was starting with a unique bidirectional LSTM layer. The following linear layer reduced channels to half of BiLSTM layer ouput channels, then we applied Rectified Linear Unit (ReLU), dropout and layer normalization. The second way was composed of one linear layer if input channel size and output channel size of the block was different, if not it returns identity. Finally, the two ways were summed and output. This residual connection allowed us to add weights to current time point data.

Two of these blocks were computed. The first one took and output hidden channel size. The second one output half of hidden size. The output block was the same as the encoding one but the first linear took half hidden size and returned quarter and the second one returned 3 vectors. Instead of the ReLU, we applied tangent hyperbolic (TanH) and then normalized in the range 0-1.

### Model Fine Tuning

We ran 366 runs with a Bayes exploration and early stopping in the range set in Supplement Table 2. This batch had a mean of minimal loss of 0.134 with standard deviation of 0.0191. The best loss achieved was 0.112. We considered the best runs as below 0.113 which represent 4 runs.

To improve the efficiency, we used a linear regression to predict the minimum value of MAE during training. The variable added to the model was the seed, learning rate, weight decay, dropout rate, gradient clip value, cosine annealing scheduler and noise maximum and minimum. The model had an adjusted R squared of 0.76 and p-value < 0.001. The seed, epoch, learning rate, noise maximum and weight decay were significant. The epoch was negatively associated with the minimal MAE and the others parameters were positively associated with MAE.

For the batch size, we predicted best value by computing mean minimal validation loss with confidence intervals. To select the optimal value of continuous parameters, we used generalized additive models (GAMs), defining optimal ranges as regions where predicted loss falls within 10% of the global minimum. Using these data, we explored a restricted hyperparameter landscape and found an optimum with the values described in the article.

### Methods Used To Decrease Overtraining

We used dropout tuning, gradient clipping, model complexity optimization, weight decay and L1/L2 regularization. These techniques were not strong enough to avoid memorization and over-fitting. We added a noise generator and tested natural and Gaussian noise. The Gaussian generator was not effective at handling overfitting. The natural noise was done by randomly permuting the batch along the first dimension. This permuted tensor was then scaled by a scheduled noise level and added to the original input. The resulting sum is normalized to get back in the original range.

### Training Method And Parameters

The training was done using a mean absolute error (MAE) loss function, AdamW optimizer^47^ and a number of channels distribution as shown in Supplement Figure 2. The learning rate was scheduled using a cosine annealing curve. The linear weight was initialized using a uniform Xavier distribution with 0.7 gain factor and the BiLSTM was initialized using a uniform Xavier distribution with 0.8 gain factor for input to hidden and Orthogonal distribution with 0.8 gain factor for hidden-to-hidden weights. To limit overfitting, we added to the signal at the batch level a natural noise described in the supplement. We compared different values of hyperparameters during training to find an optimum detailed in Supplement Table 2. We tuned batch size, weight decay, dropout rate, learning rate, learning rate scheduler parameter, gradient clip value, seed, and noise scheduling parameters.

After tuning, final parameters were learning rate set at 0.0005 with a cosine period of 5 without modification of the period though epochs, a batch size of 32, a weight decay of 0.2, a gradient clip value of 0.8, seed of 123, and noise of 0.5 to 0 with a linear decrease from epoch 2 to 200.

### Model Feature Importance

The model’s feature importance was tested using SHapley Additive exPlanations (SHAP) values using the DeepExplainer algorithm^48^. We computed SHAP values for the prediction of the last timepoints of each block through particpants. Background data was set using the training dataset and SHAP values were computed with validation and test sets. This method allowed us to have a feature importance for each participant, each timepoint, and each ROI. In order to have the SHAP value for each of the 18 networks described, we L2 regularized the network vector and SHAP value vector. After L2 regularization, each reduction of the shape of the matrix was done using a min-max scaled mean absolute value.

## Acknowledgements

We would like to thank Dr. Meryl Butters for her feedback on this manuscript. Research reported in this publication was supported by the National Institute of Mental Health of the National Institutes of Health under award number R01MH108509, K01 MH122741, R21 MH138955, R01 MH 121619, T32 MH019986.

## Data availability

The fMRI data that support the findings are available from the corresponding authors upon reasonable request.

## Code availability

Code is accessible via GitHub: https://github.com/tetra-tools/JYC-HK-wfMRIWorryPrediction/tree/main.

## Contributions

Study Conception and Design – JYC, HTK, CA; Data Acquisition – HTK, CA; Analysis and Interpretation of Data – JYC, HTK, CA; Drafting – JYC, HTK, CA; and Critical Revision – JYC, HTK, CA, TD, DT

## Supplementary

**Supplement Table 1.** Population Description.

| Variable | N | FINA Train | FINA Validation | RAW |
| --- | --- | --- | --- | --- |
|  |  | N = 92<sup>1</sup> | N = 24<sup>1</sup> | N = 88<sup>1</sup> |
| **Age** | 203 | 59 (54, 64) | 62 (55, 70) | 62 (56, 69) |
| Unknown |  | 1 | 0 | 0 |
| **Sex** | 204 |  |  |  |
| Male |  | 35 (38%) | 8 (33%) | 15 (17%) |
| Female |  | 57 (62%) | 16 (67%) | 72 (82%) |
| Others |  | 0 (0%) | 0 (0%) | 1 (1.1%) |
| **Education** | 183 | 16.00 (14.00, 18.00) | 16.00 (14.00, 18.00) | 16.00 (16.00, 18.00) |
| Unknown |  | 6 | 3 | 12 |
| **PSWQ** | 204 | 50 (40, 65) | 49 (36, 61) | 41 (32, 56) |
| **RSQ** | 197 | 37 (29, 45) | 33 (27, 47) | 30 (27, 40) |
| Unknown |  | 7 | 0 | 0 |
| **HARS** | 201 | 8.0 (4.0, 12.0) | 6.5 (2.5, 12.5) | 7.0 (3.0, 12.0) |
| Unknown |  | 3 | 0 | 0 |
| **HAMD** | 88 |  |  | 5 (2, 6) |
| Unknown |  |  |  | 0 |
| **MADRS** | 109 | 7 (2, 14) | 4 (2, 12) |  |
| Unknown |  | 6 | 1 |  |

**Supplement Figure 1.**
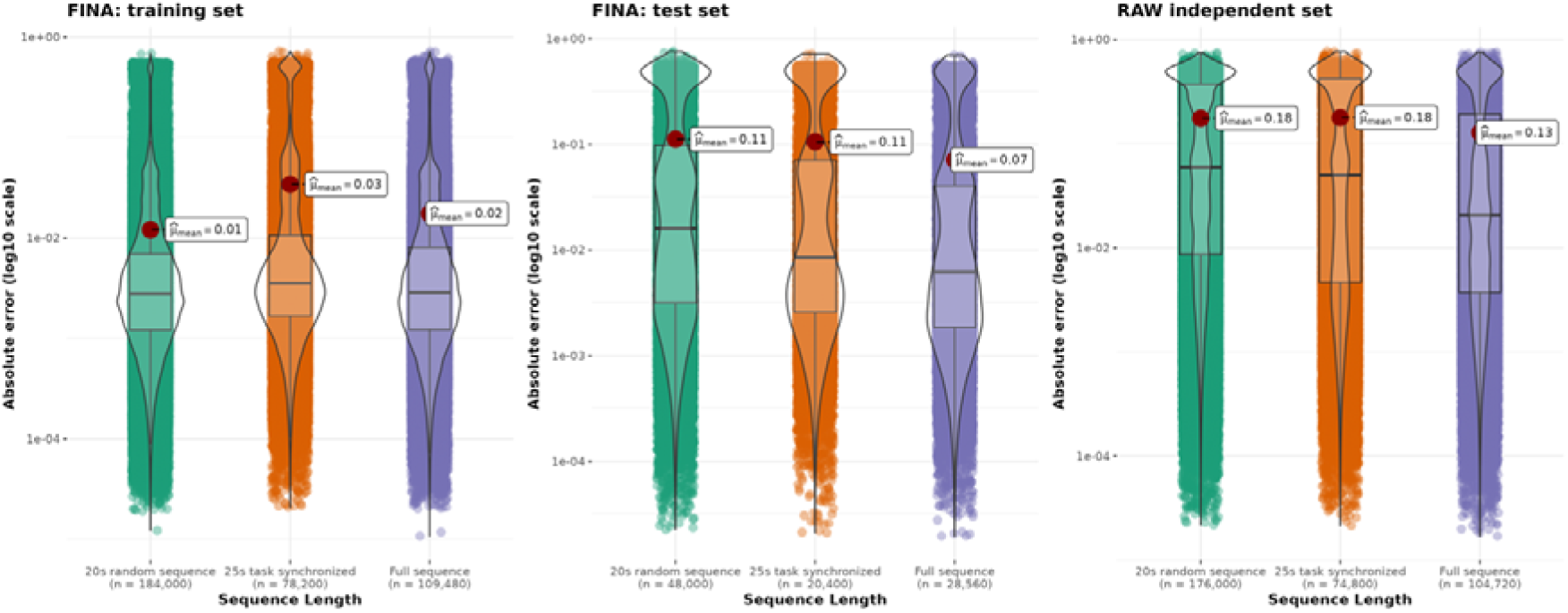
Model performance for each dataset expressed as absolute error for different sequence length and each subset. Absolute error distribution showed for each extracted sequence length and dataset as a violin plot. This absolute error is expressed as log^10^ scaled axis. A boxplot is added to show the median and quartiles. The red dot shows the mean value. The sequence length label shows the number of timepoints used to compute each distribution (100 x 20s random sequence, 34 x 25s task synchronized, and 1190s full sequence per participant). This figure underline the small effect of sequence length. The **FINA: training set** has predictions around 0.02 +/- 0.01 mean absolute error compared to 0.07-0.11 mean absolute error in **FINA: test set** and 0.13-0.18 mean absolute error in **RAW independent set**.

**Supplement Table 2.**
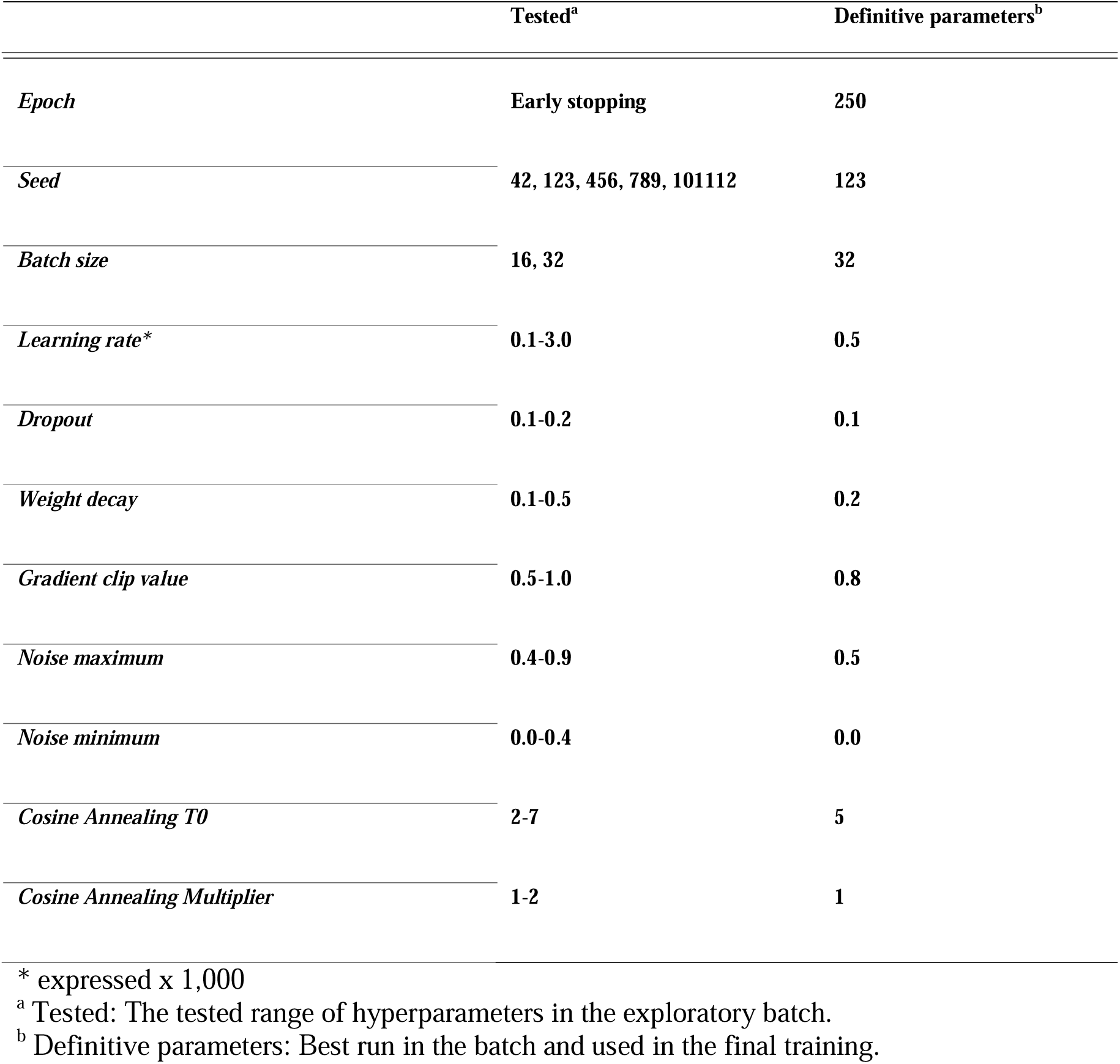
Hyperparameters exploration and best set of hyperparameters.

**Supplement Figure 2.**
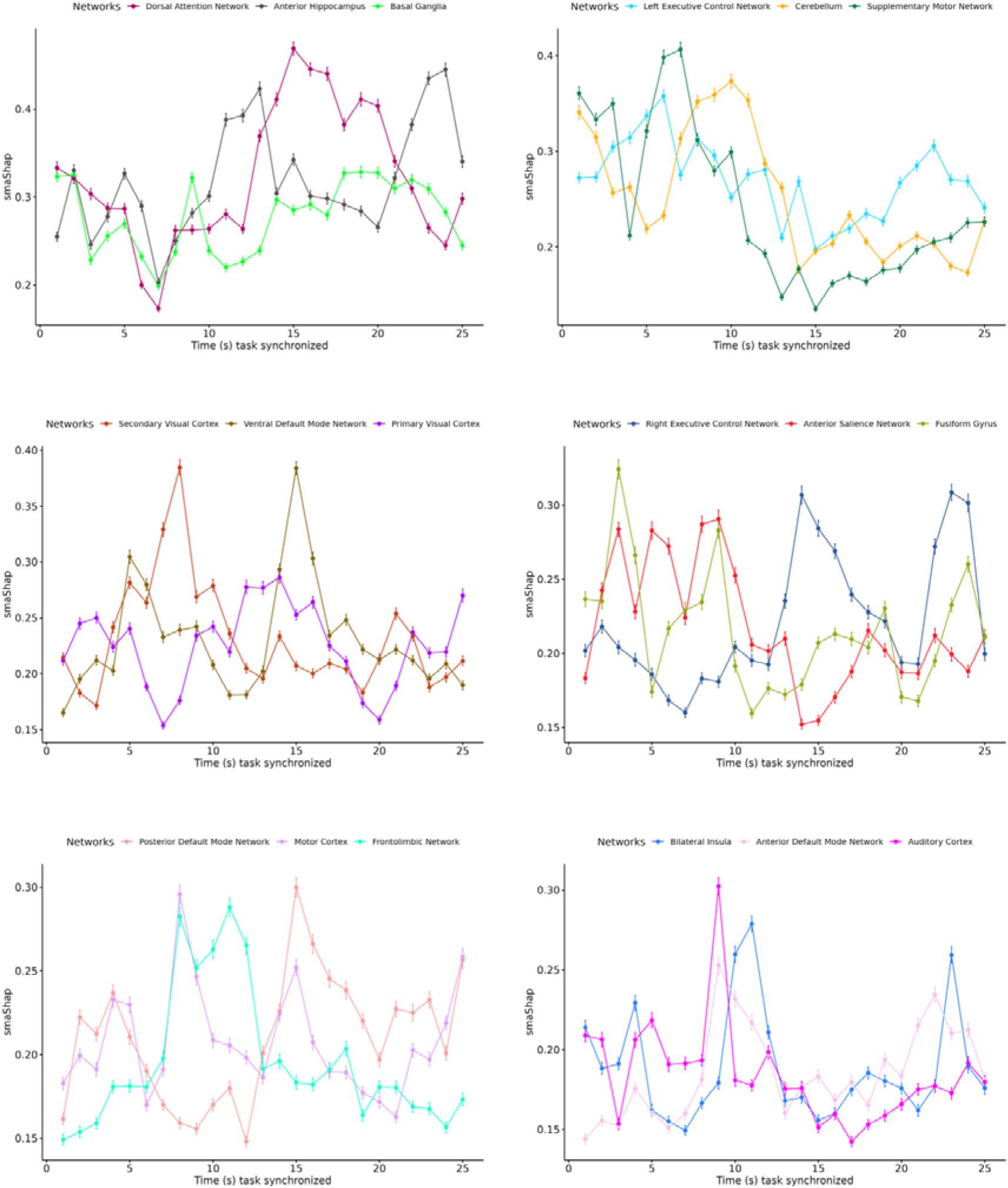
Worry smaShap dynamics. Scaled mean absolute of Shap values (smaShap) of all networks are represented for worry. Networks are in descending order based on the 25s mean of smaShap. At each timepoint, the networks smaShap values are reported by their means and 95% confidence intervals.

**Supplement Figure 3.**
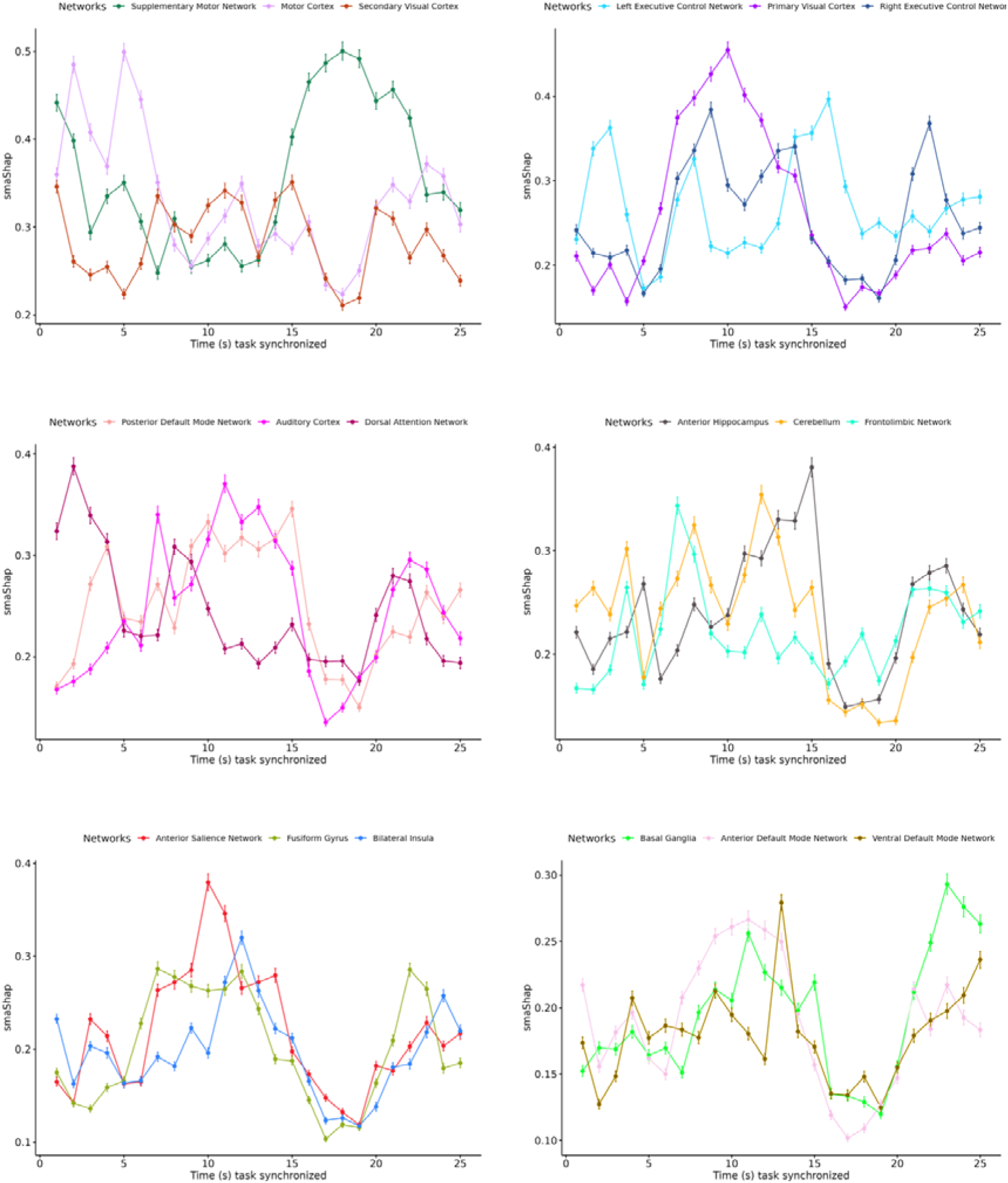
Neutral smaShap dynamics. Scaled mean absolute of Shap values (smaShap) of all networks are represented for neutral. Networks are in descending order based on the 25s mean of smaShap. At each timepoint, the networks smaShap values are reported by their means and 95% confidence interval.

**Supplement Figure 4.**
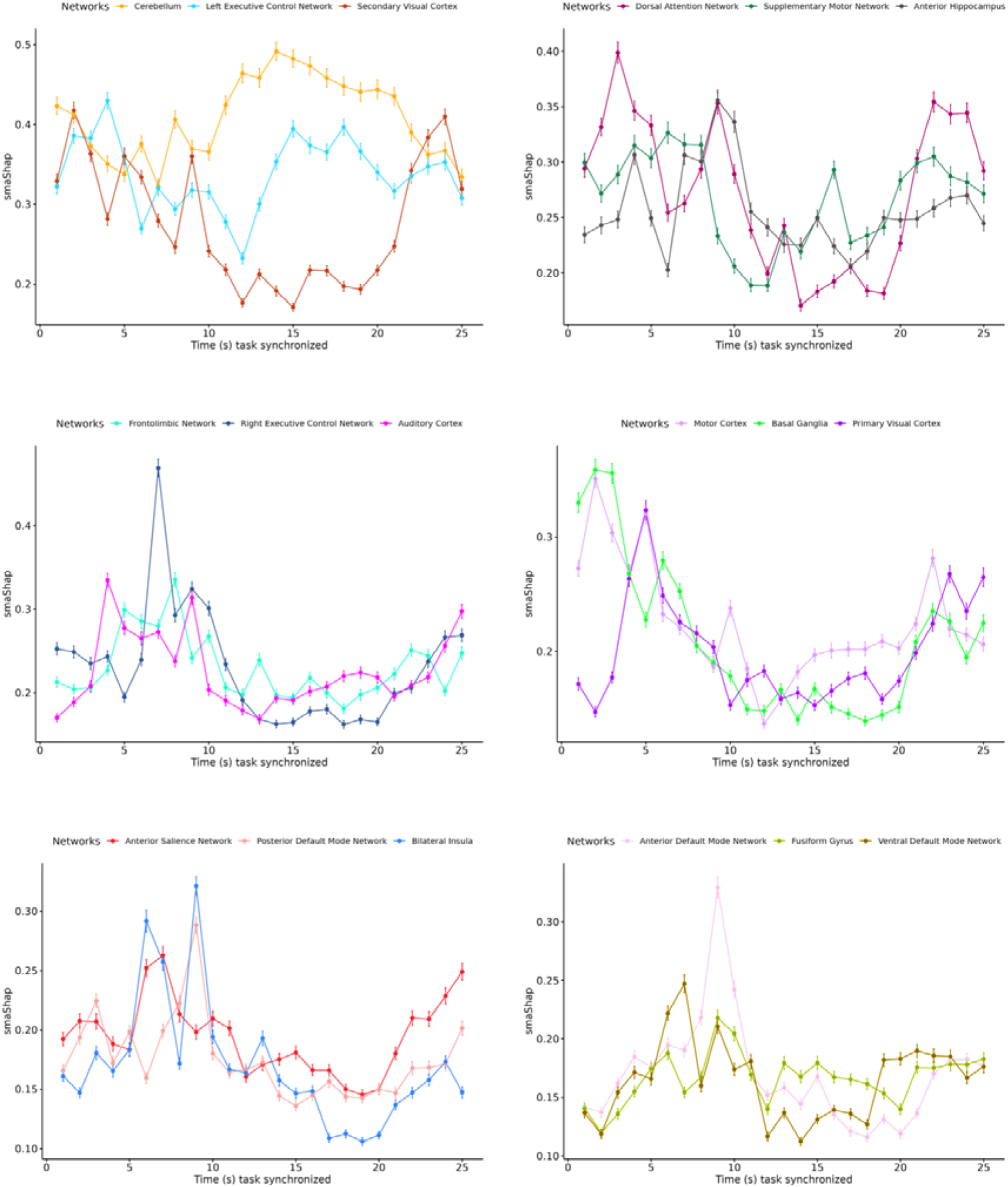
Reappraisal smaShap dynamics. Scaled mean absolute of Shap values (smaShap) of all networks are represented for reappraisal. Networks are in descending order based on the 25s mean of smaShap. At each timepoint, the networks smaShap values are reported by their means and 95% confidence intervals.

**Supplement Figure 5.**
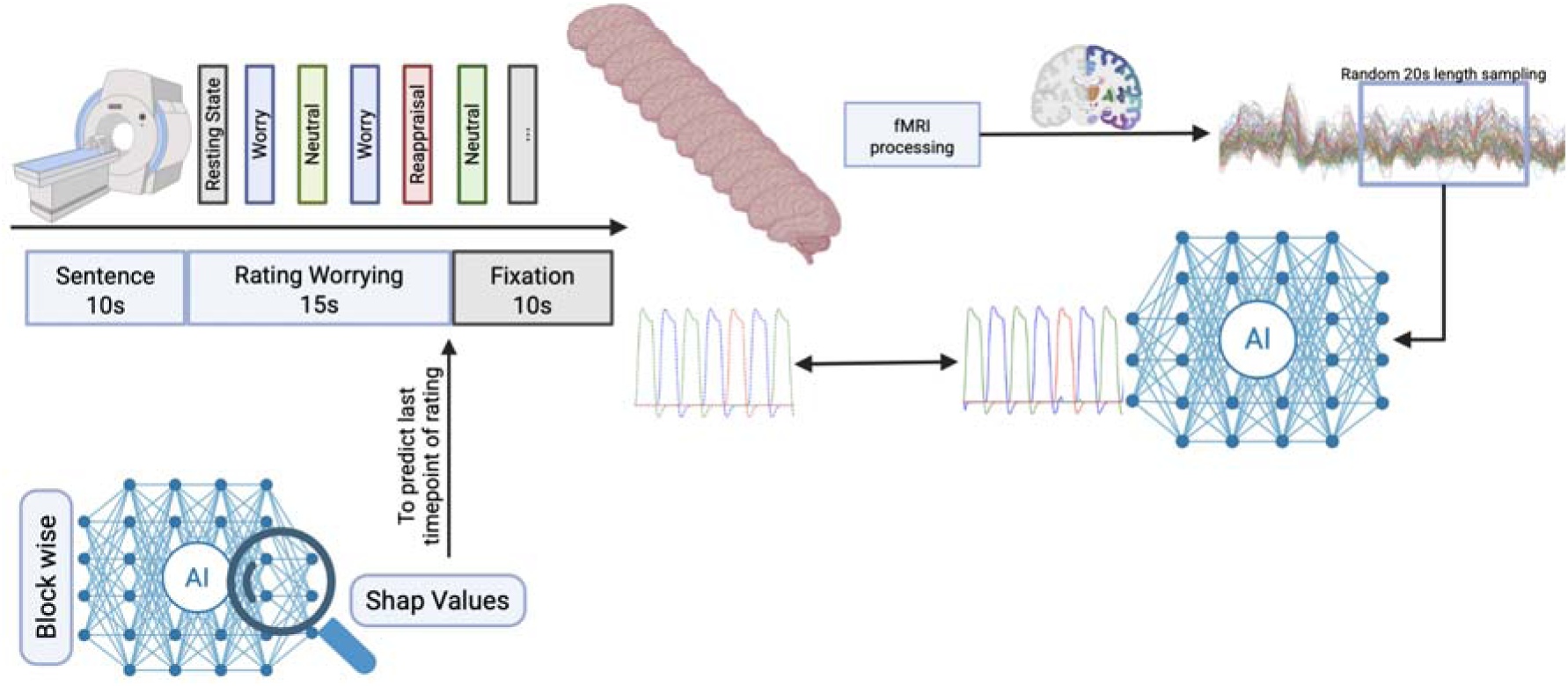
Experimental design. This representation illustrate the experimental design. The fMRI task is designed with a leading 10 second of resting state followed by 16 block of worry separated by a neutral and/or a reappraisal block. Each block is composed by a 10 seconds sentence exposition, 15 seconds of rating and 10 seconds of cross fixation. Each fMRI is preprocessed and segmented into regions of interest and these signals are randomly sampled into 20 seconds length sequences. The model is predicting the actual block class activation and compared for training. Shap values are predicted using a block wise sampling of the signal.

**Supplement Figure 6.**
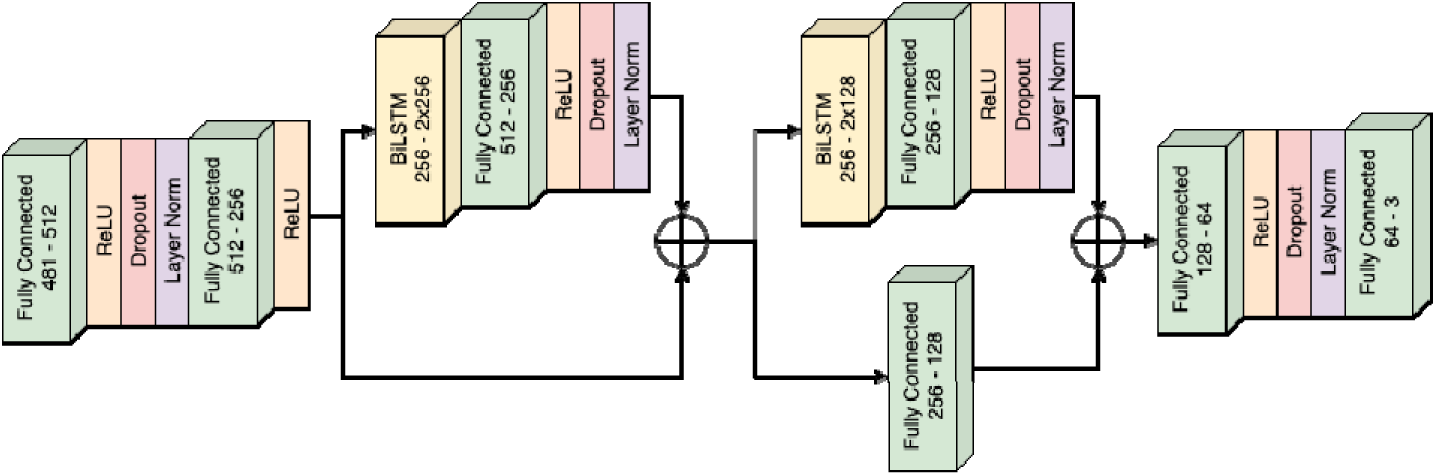
Model design. This figure represent the final model architecture. Fully connected layers are represented in green with a label specifying the size of input channels and output channels. The Rectified Linear Unit is represented in orange. The dropout is represented in red with a respective dropout of 0.05, 0.1, 0.1, 0.025 from left to right. Layer normalization is perform over the channel dimension and colored in purple. Arrows shows the flow of data and the cross inside a circle represents an addition of the data coming in. The data always have the same dimensional size when an addition is applied.

